# The association between enlarged perivascular spaces and muscle sympathetic nerve activity in normotensive and hypertensive humans

**DOI:** 10.1101/2024.12.23.24319592

**Authors:** Donggyu Rim, William Pham, Rania Fatouleh, Annemarie Hennessy, Markus Schlaich, Luke A Henderson, Vaughan G Macefield

## Abstract

**Background:** Perivascular spaces (PVS) are fluid-filled spaces that functions as channels for glymphatic clearance in the brain. Enlarged perivascular spaces (ePVS) have been associated with high blood pressure. Hypertension features abnormal increases in muscle sympathetic nerve activity (MSNA), which constricts blood vessels in the muscle vascular bed, but the underlying mechanisms for this increase are not understood. Moreover, the association between ePVS and the resting sympathetic outflow from the brain has not been studied in normotensive or hypertensive humans. Therefore, we assessed whether ePVS is associated with muscle sympathetic nerve activity (MSNA) in 25 hypertensive patients and 50 healthy normotensive adults.

**Methods:** T1-weighted MRI anatomical brain images were analysed for ePVS using a deep learning-based segmentation algorithm – nnU-Net. ePVS in the white matter (WM), basal ganglia (BG), hippocampus (HP), and midbrain (MB) were analysed. Spontaneous bursts of MSNA were recorded from the right common peroneal nerve via a tungsten microelectrode immediately before the MRI scan.

**Results:** Significant associations were found between ePVS and MSNA in the WM, BG, and HP in both the normotensive and hypertensive groups after adjusting for confounding factors (age, sex, mean blood pressure, total intracranial volume). However, the association between MSNA and MB ePVS was only observed in the hypertensive group.

**Conclusion:** This finding provides insights into the pathophysiology of elevated sympathetic drive in hypertension.

*What is new?:* • Enlarged perivascular spaces (ePVS) are associated with muscle sympathetic nerve activity (MSNA) in normotensive and hypertensive humans. • Hypertensives and normotensives display differences in the association between the midbrain ePVS and MSNA.

*What are the clinical implications?:* • Insights into the centrally driven pathophysiological mechanism of elevated sympathetic nerve outflow in hypertension has been revealed. • ePVS may be used as an imaging biomarker for individuals with high sympathetic nerve activity, allowing identification of risk individuals that require microneurographic assessment of sympathetic nerve activity

## Introduction

Perivascular spaces (PVS) are fluid-filled spaces surrounding cerebral blood vessels, formed between the vascular smooth muscle basement membrane and the ensheathing astrocyte end- feet ^1^. In disease, PVS can become wider, and hence, visible on T1-weighted magnetic resonance imaging (MRI), known as enlarged PVS (ePVS). ePVS is associated with hypertension ^2–4^ as well as with neurodegenerative diseases such as Parkinson’s Disease and Alzheimer’s Disease ^5,6^ and is associated with increased risk of vascular events, including myocardial infarction and stroke ^7,8^, as well as hypertension. It is believed that the ePVS may contribute to neuroinflammation ^9^ and represent disrupted waste clearance through the glymphatic system ^10^.

Muscle sympathetic nerve activity (MSNA) is generated in the brain and contributes to the beat-to-beat regulation of blood pressure. In humans, it can be recorded by intraneural microelectrodes inserted into peripheral nerves and could serve as a physiological marker of the brain’s vasoconstrictor drive. Indeed, we know that sympathetic outflow is influenced by neuroinflammation ^11–13^, astrocytes ^14–17^, and brain structure ^18,19^. Given that these factors may also contribute to dysregulated perivascular spaces, it is not unreasonable to think that it may also contribute to the abnormal increases in sympathetic outflow associated with hypertension and the elevated risk of stroke ^20,21^. However, despite the damaging effect of abnormal sympathetic nerve activity elevations, the underlying mechanism of increased sympathetic nerve activity in hypertension is still poorly understood.

In this study, we aimed to assess whether sympathetic nerve activity is associated with MRI-visible ePVS in both normotensive and hypertensive adults. There are increasing evidence suggesting that structural and functional changes in the brain could be associated with changes in sympathetic outflow ^19^. We have shown that normotensive participants with higher grey matter volume in the dorsomedial hypothalamus (DMH) and posterior cingulate cortex had lower MSNA ^18^. Also, in patients with obstructive sleep apnoea (OSA), regions that are functionally coupled to the generation of bursts of MSNA showed augmented blood oxygen level-dependent (BOLD) signals in functional MRI compared to age-matched controls ^22^. These studies provide evidence that structural and functional brain changes could lead to altered sympathetic outflow to peripheral organs. We hypothesised that perivascular spaces may also influence sympathetic outflow. Moreover, given that PVS can be identified in standard T1-weighted clinical MRI scans, measurement of PVS may be a method to monitor early pathophysiological changes in sympathetic outflow that may lead to increases in blood pressure.

## Methods

### Study design

This cross-sectional study was approved by the Western Sydney University Human Research Ethics Committee (HREC approval H11462) and endorsed by Governance at the Baker Heart and Diabetes Institute. Informed written consent was obtained from all participants and conformed to the Declaration of Helsinki. All participants refrained from caffeinated drinks and nicotine on the morning of the experiment until completion. All experiments were conducted at 12 PM.

### Participants

25 hypertensive patients and 50 healthy normotensive participants were recruited for this study. Participants previously diagnosed with hypertension and currently using peripherally acting antihypertensive medications, and participants that had a seated average systolic BP

≥130 mmHg and diastolic BP ≥80 mmHg were classified as hypertensive for this study. Participants were excluded if they (i) had a previous history of cardiovascular disease such as heart failure or stroke; (ii) were taking antihypertensive medications with centrally acting mechanisms such as moxonodine or methyldopa; (iii) had conditions contraindicated for MRI scanning, such as having cardiac pacemakers, metallic implants, or aneurysm clips.

### Data acquisition

Blood pressure measurement was taken after a 5-minute resting period in a seated position with an automated sphygmomanometer (UA-651SL, A&D, Japan). An average of three blood pressure measurements, taken with a 1-minute rest period in between the measurements, was reported.

Microneurography was performed with participants laying supine on an MRI bed. An insulated tungsten microelectrode (200μm, Frederick Haer and Co, Bowdoin, ME, United States) was inserted into the common peroneal nerve of the right leg, with an uninsulated reference microelectrode inserted ∼1 cm away, at a depth of 1-2 mm. Raw signals detected with microelectrodes were amplified (NeuroAmp EX, ADInstruments, Sydney, Australia) to 2 x 10^4^, with bandpass of 0.3-5.0 kHz. Signals were stored on a computer data acquisition and analysis system (PowerLab 16SP hardware and LabChart for Macintosh, v7.2.5; ADInstruments). To map the location of the common peroneal nerve, a small weak transcutaneous electrical stimulation (0.2ms, 1-10mA, 1Hz) was delivered near the fibular head via an external probe. The site of percutaneous insertion of an insulated microelectrode was determined by the locations that evoked the strongest twitches with the external probe. The insertion of the microelectrode tip in the muscle fascicle was confirmed by a twitch of muscles acting on the foot (tibialis anterior, extensor digitorum longus, or peroneus longus) at stimulus currents of 0.02 mA or less. The identity of the muscle fascicle was confirmed by the presence of an afferent activity of muscle spindles during passive stretching of the parent muscle, as well as the absence of afferent impulses evoked by stroking the skin of the fascicular innervation territory of the common peroneal nerve. Further manual manipulations of the microelectrode were done to search for spontaneous bursts of muscle sympathetic nerve activity (MSNA) that were identified by having characteristics of strong cardiac rhythmicity as well as an increased activity during a maximal inspiratory breath hold but an absence of response during unexpected arousal stimuli. Raw MSNA signals were processed as a Root Mean Squared (RMS, moving average 200 ms) signal for enhanced visualisation of the bursts. In addition to microneurography, electrocardiogram (ECG, 2kHz, 3 Ag/AgCl surface electrodes (Covidien, Ireland)), and continuous non-invasive blood pressure recording were obtained from finger cuffs (NOVA, Finapres Medical System BV, Netherlands). MSNA bursts were counted using the Cyclic Measurements peak detection feature of LabChart. The number of bursts in one minute (burst frequency, BF), and 100 heartbeats (burst incidence, BI) were calculated for each participant. More detailed recording and quantification methods of MSNA are outlined in our review (Macefield et al., 2021).

### Imaging acquisition

Thirty-nine T1-weighted anatomical images of the brain were acquired using a Siemens Magnetom 3T (64-channel SENSE coil, repetition time [TR]=2300ms, echo time [TE]=2.49ms, flip angle=8°, 192 sagittal slices, voxel size=0.9mm isotropic), while 59 T1- weighted anatomical scans were obtained using a Philips Achieva 3T (32-channel SENSE head coil TR=5600 ms, TE=2.5ms, flip angle=8°, 200 sagittal slices, voxel size=0.87mm isotropic).

### Imaging processing

Enlarged perivascular spaces (ePVS) were identified using a deep learning-based segmentation algorithm (nnU-Net residual encoder) from the T1-weighted anatomical scans. nnU-net is a self-configuring deep-learning-based segmentation method that incorporates preprocessing, network architecture, training, and post-processing for biomedical images (Isensee et al., 2021). The U-Net architecture relies on convolutional filters arranged in a U-shaped configuration, to extract image features of interest. Upon visual inspection, the model can accurately segment PVS voxels in white matter, basal ganglia, hippocampus, and midbrain. Subsequently, it was used to predict PVS in this dataset (n=75) as shown in Fig. 1. All identified MRI-PVS were considered enlarged as the model was trained with manually labelled enlarged perivascular spaces (ePVS) reviewed by a radiologist. The total number of distinct ePVS voxel clusters was represented as “ePVS cluster”, and the total PVS voxels in all clusters multiplied by the voxel size of the image was represented as “ePVS volume”. The algorithm was further optimised by using the sparse annotation method, target spacing optimisation, image preprocessing optimisation using non-local means filtering (NLMF) and adaptive histogram equalisation (AHE), and semi-supervised learning for T1-weighted anatomical images ^23^. Total intracranial volume (TIV) was obtained with a computational anatomy toolbox (CAT12) ^24^ for statistical parametric mapping (SPM) version 12.

**Figure 1.**
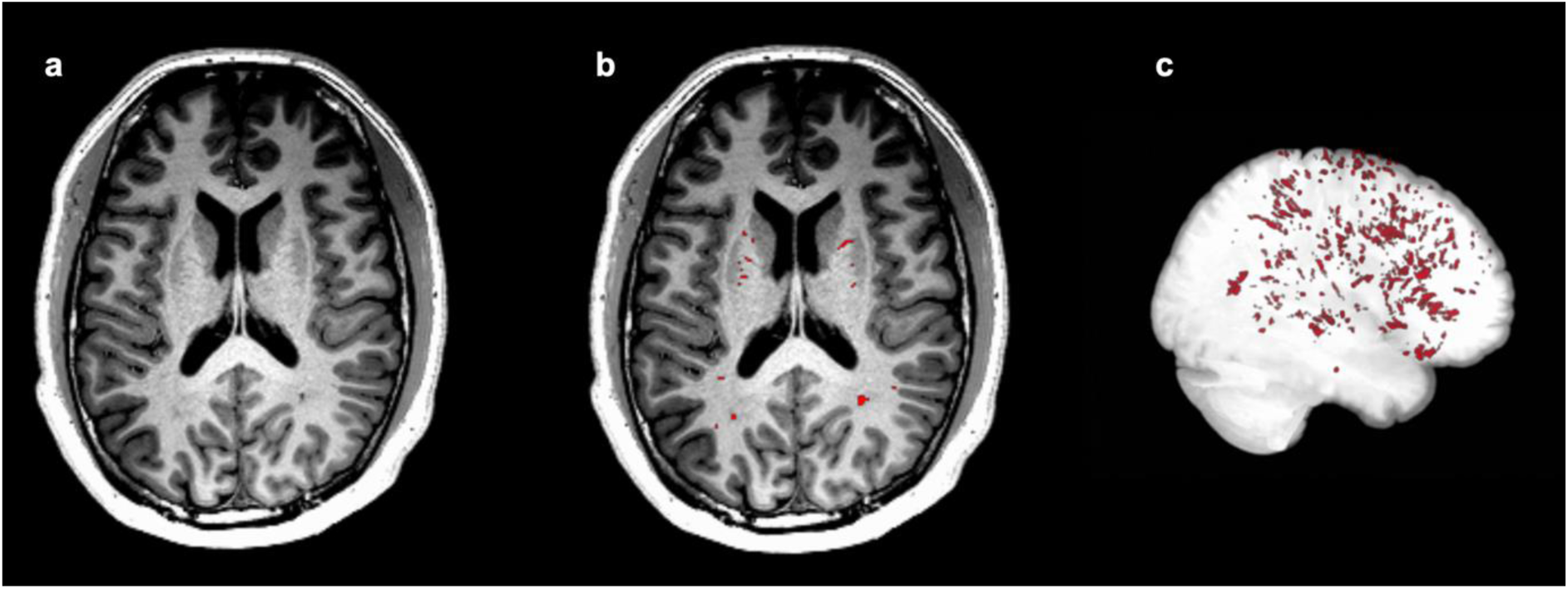
Perivascular spaces segmentation example using nnU-Net, deep learning-based segmentation methods. **a** axial slice of T1- weighted MRI before segmentation. **b** representation of segmented perivascular spaces on the T1-weighted MRI indicated by red. Labelling of perivascular spaces appears in the basal ganglia and white matter regions. **c** 3D-rendered perivascular spaces segmentation across the whole brain

### Statistical analysis

Statistical analyses were performed using GraphPad Prism (v9.5.1 for Macintosh, San Diego, CA, USA) and IBM SPSS Statistics (v26.0, Armonk, NY, USA). Participant characteristics were assessed for normality using the Shapiro-Wilk test. Continuous variables are reported as mean ± standard error of the means as well as 95% CI interval. Categorical variables are shown as percentages. We conducted a correlation analysis using a Spearman nonparametric correlation matrix with GraphPad Prism to determine the association between ePVS (count or volume) and MSNA (incidence or frequency) as well as other participant characteristics such as age, sex, and haemodynamic measurements. For the ePVS regions that showed a significant correlation with MSNA, additional group-level simple linear regressions were conducted between the ePVS and each MSNA metric (BI and BF). A backward multiple regression analysis was conducted to assess the relationship between ePVS and MSNA after adjusting for confounding factors such as age, sex, mean blood pressure and TIV. Moreover, additional backward multiple regression analysis was conducted to assess the relationship between ePVS and haemodynamic measurements such as the mean blood pressure. The criterion for the backward elimination method was set at a probability of F-to-remove greater than or equal to 0.1.

## Results

A total of 75 participants were included in the study. Twenty-five participants were hypertensive patients, and 50 participants were normotensive. The two groups had no statistically significant differences in age. However, significant differences were observed in the male-to-female ratio between the groups. As expected, all haemodynamic measurements were elevated in the hypertension group compared to the normotensive group. The standard metrics of muscle sympathetic nerve activity measurements were elevated in the hypertensive group compared to the controls, reflecting the increases in central sympathetic drive to the peripheral blood vessels with hypertension. Overall, the number of white matter ePVS in the hypertension group was reduced compared to the normotensive group, but the volume of white matter ePVS was not significantly different between the groups. Conversely, there were no statistically significant differences in ePVS between the two groups for the basal ganglia, hippocampus, and midbrain. There was no statistically significant difference in the total intracranial volume (TIV) between the groups, but the hypertensive group had reduced total grey matter volume and total white matter volume. Data are presented in Table 1.

**Table 1.**
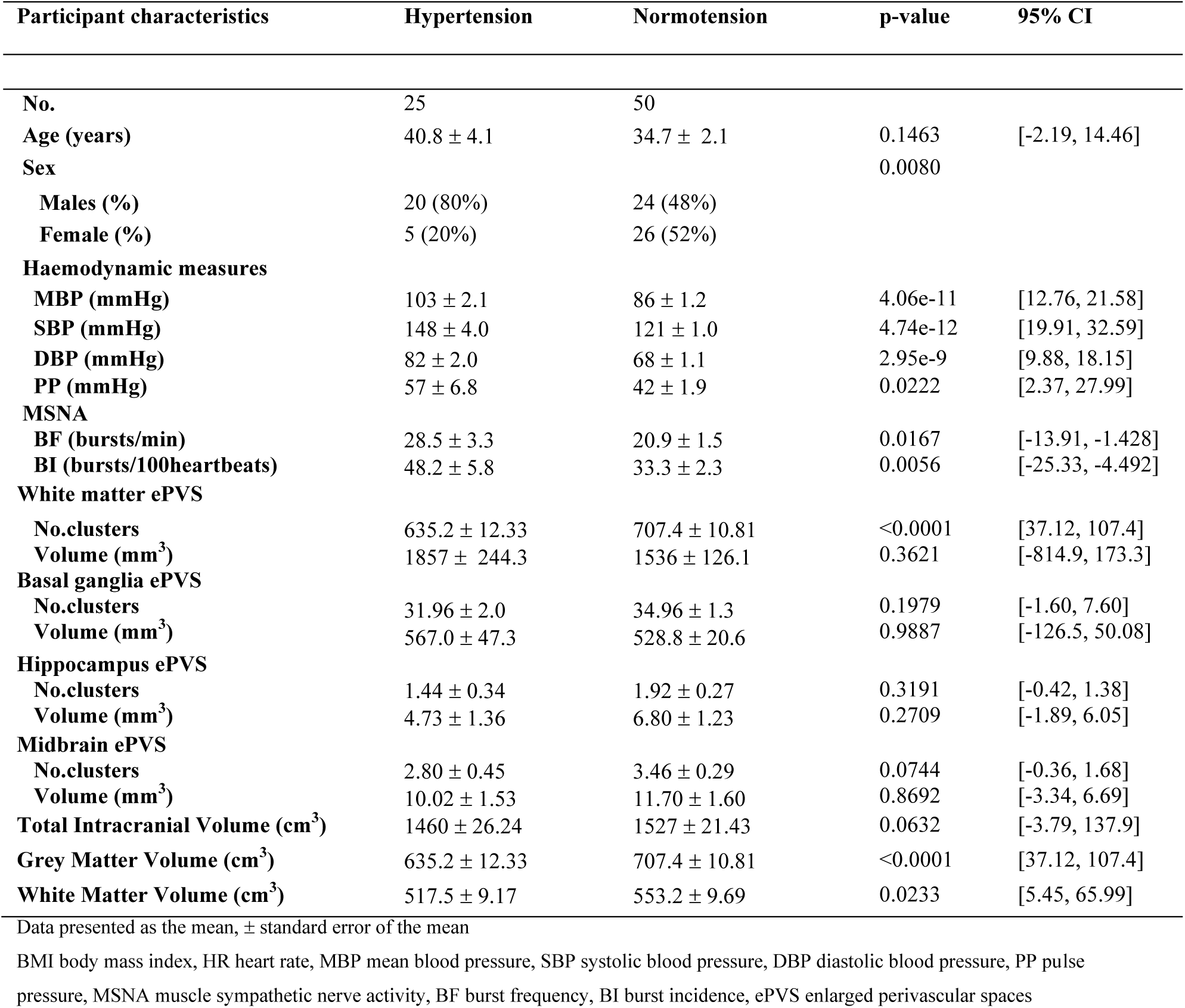
Participant characteristics

| Participant characteristics | Hypertension | Normotension | p-value | 95% CI |
| --- | --- | --- | --- | --- |
| **No.** | 25 | 50 |  |  |
| **Age (years)** | 40.8 ± 4.1 | 34.7 ± 2.1 | 0.1463 | [-2.19, 14.46] |
| **Sex** |  |  | 0.0080 |  |
| **Males (%)** | 20 (80%) | 24 (48%) |  |  |
| **Female (%)** | 5 (20%) | 26 (52%) |  |  |
| **Haemodynamic measures** |  |  |  |  |
| **MBP (mmHg)** | 103 ± 2.1 | 86 ± 1.2 | 4.06e-11 | [12.76, 21.58] |
| **SBP (mmHg)** | 148 ± 4.0 | 121 ± 1.0 | 4.74e-12 | [19.91, 32.59] |
| **DBP (mmHg)** | 82 ± 2.0 | 68 ± 1.1 | 2.95e-9 | [9.88, 18.15] |
| **PP (mmHg)** | 57 ± 6.8 | 42 ± 1.9 | 0.0222 | [2.37, 27.99] |
| **MSNA** |  |  |  |  |
| **BF (bursts/min)** | 28.5 ± 3.3 | 20.9 ± 1.5 | 0.0167 | [-13.91, -1.428] |
| **BI (bursts/100heartbeats)** | 48.2 ± 5.8 | 33.3 ± 2.3 | 0.0056 | [-25.33, -4.492] |
| **White matter ePVS** |  |  |  |  |
| **No.clusters** | 635.2 ± 12.33 | 707.4 ± 10.81 | <0.0001 | [37.12, 107.4] |
| **Volume (mm<sup>3</sup>)** | 1857 ± 244.3 | 1536 ± 126.1 | 0.3621 | [-814.9, 173.3] |
| **Basal ganglia ePVS** |  |  |  |  |
| **No.clusters** | 31.96 ± 2.0 | 34.96 ± 1.3 | 0.1979 | [-1.60, 7.60] |
| **Volume (mm<sup>3</sup>)** | 567.0 ± 47.3 | 528.8 ± 20.6 | 0.9887 | [-126.5, 50.08] |
| **Hippocampus ePVS** |  |  |  |  |
| **No.clusters** | 1.44 ± 0.34 | 1.92 ± 0.27 | 0.3191 | [-0.42, 1.38] |
| **Volume (mm<sup>3</sup>)** | 4.73 ± 1.36 | 6.80 ± 1.23 | 0.2709 | [-1.89, 6.05] |
| **Midbrain ePVS** |  |  |  |  |
| **No.clusters** | 2.80 ± 0.45 | 3.46 ± 0.29 | 0.0744 | [-0.36, 1.68] |
| **Volume (mm<sup>3</sup>)** | 10.02 ± 1.53 | 11.70 ± 1.60 | 0.8692 | [-3.34, 6.69] |
| **Total Intracranial Volume (cm<sup>3</sup>)** | 1460 ± 26.24 | 1527 ± 21.43 | 0.0632 | [-3.79, 137.9] |
| **Grey Matter Volume (cm<sup>3</sup>)** | 635.2 ± 12.33 | 707.4 ± 10.81 | <0.0001 | [37.12, 107.4] |
| **White Matter Volume (cm<sup>3</sup>)** | 517.5 ± 9.17 | 553.2 ± 9.69 | 0.0233 | [5.45, 65.99] |

Data presented as the mean, ± standard error of the mean
BMI body mass index, HR heart rate, MBP mean blood pressure, SBP systolic blood pressure, DBP diastolic blood pressure, PP pulse pressure, MSNA muscle sympathetic nerve activity, BF burst frequency, BI burst incidence, ePVS enlarged perivascular spaces

The association between MSNA with ePVS, and its relationships with other participant characteristics were first assessed using a Spearman correlation matrix (Fig. 2). MSNA BF and BI were positively associated with ePVS volume in the basal ganglia (BG), as well as with midbrain (MB). Interestingly, the basal ganglia, midbrain, and white matter ePVS volumes were positively associated with age. Moreover, MSNA BF and BI showed a positive association with age. With regards to the haemodynamic measures, MSNA BI was positively correlated with mean blood pressure (MBP) as well as with diastolic blood pressure (DBP) but was negatively correlated with pulse pressure (PP). MSNA BF was not associated with MBP but was positively associated with DBP and negatively associated with PP. The numbers of ePVS clusters in the BG and MB showed negative correlations with DBP. WM ePVS cluster was positively correlated with PP and sex.

**Figure 2.**
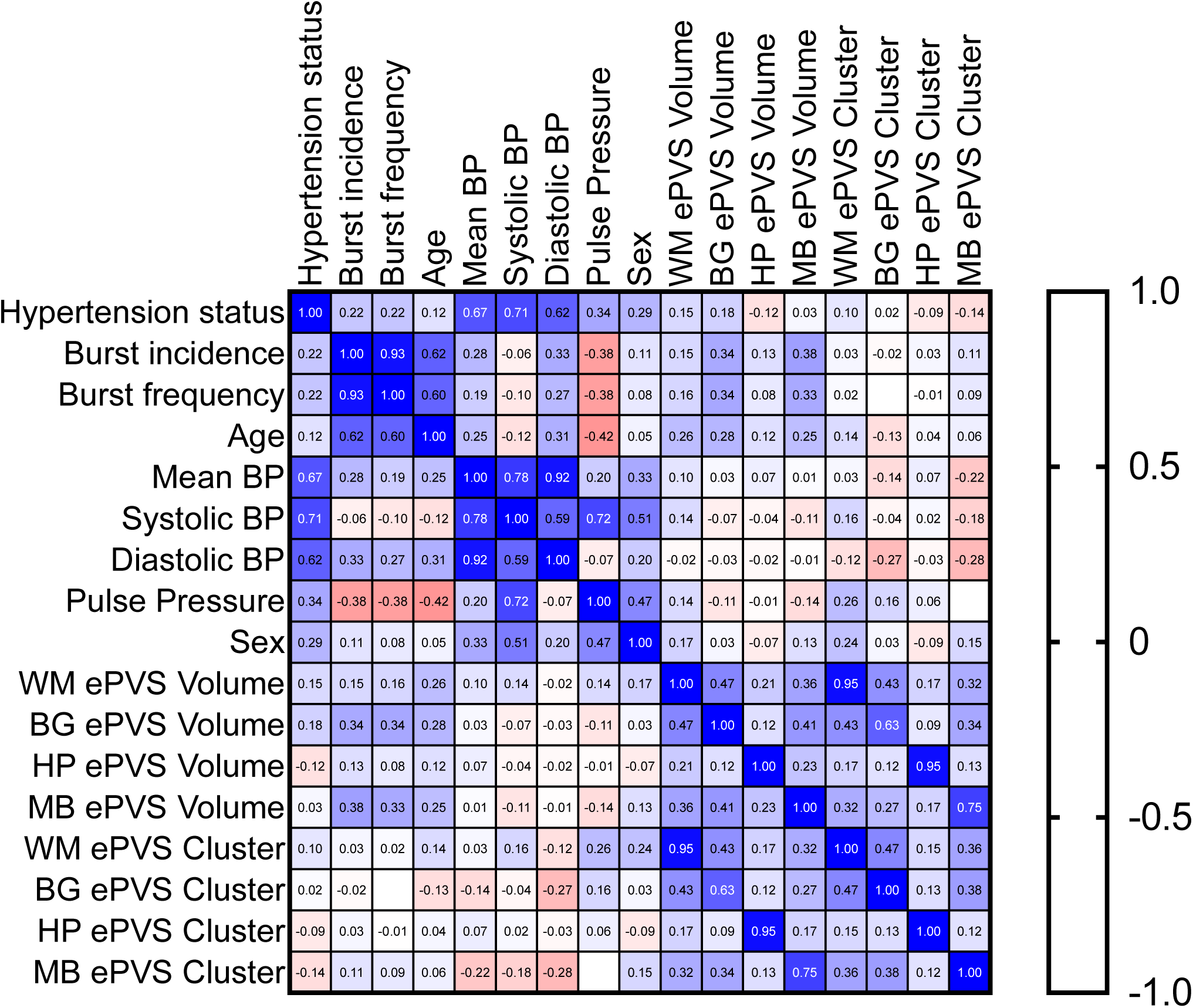
Correlation matrix showing associations of MSNA, PVS, Age, Haemodynamic measures, and sex. n=75 (pooled data from: 25 hypertensives and 50 normotensive controls) PVS perivascular space BP blood pressure WM white matter BG basal ganglia HCP hippocampus MB midbrain. Spearman Correlation coefficient values are shown. WM ePVS was adjusted with total WM value. BG, HCP, and MB ePVS was adjusted with total GM value

In the brain region that showed a significant correlation between MSNA and ePVS volume, the group-level associations between MSNA and ePVS volume were assessed with simple linear regression analysis (Fig. 3). We observed a significant positive correlation between MSNA BF and BI with BG ePVS volume in both the normotensive group and the hypertensive group. Interestingly, the correlation between MSNA BF and BI with midbrain ePVS volume was only observed in the normotensive group. No correlation was observed between WM or HP ePVS and MSNA.

**Figure 3.**
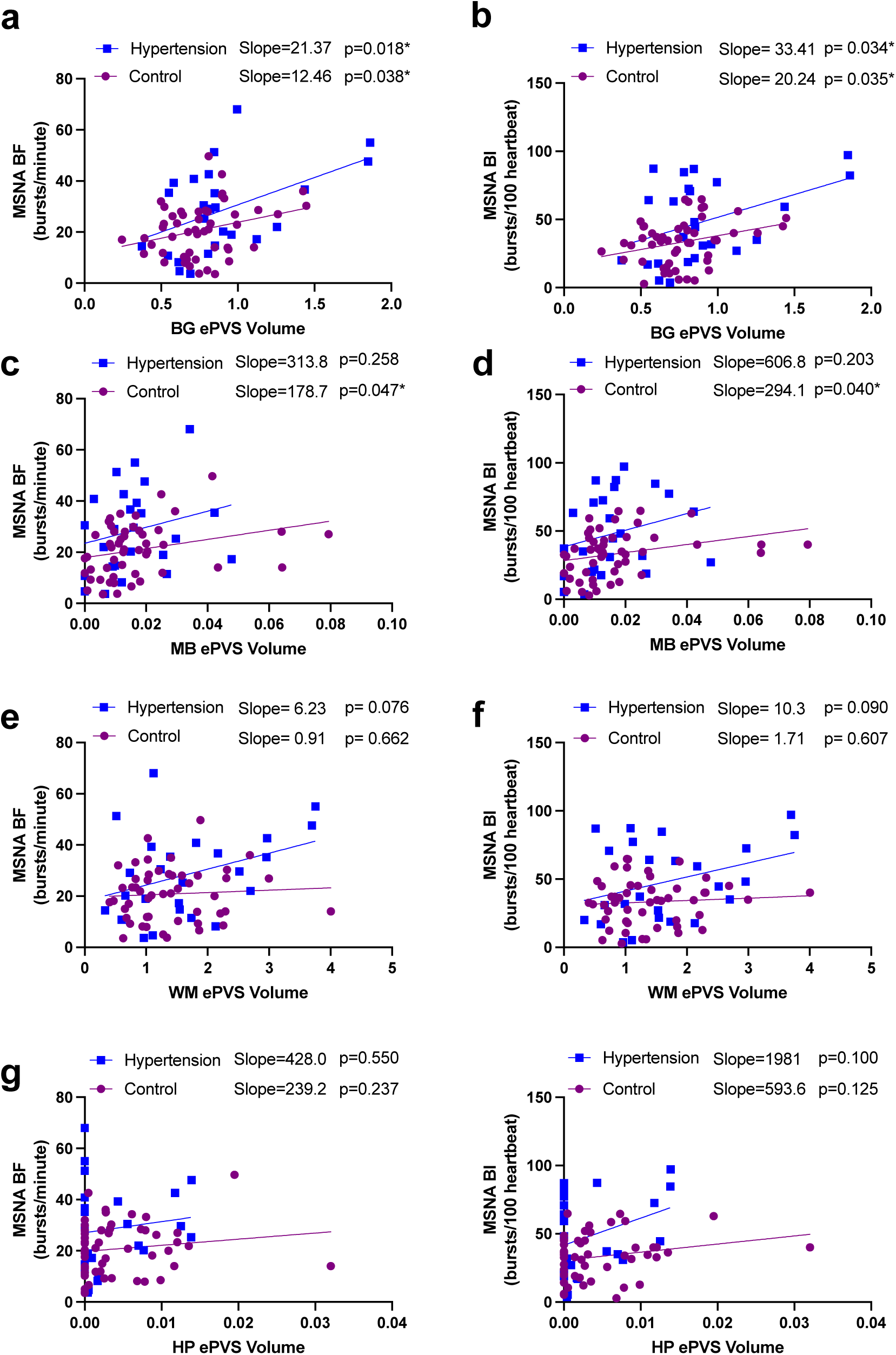
Simple linear regression between BG and MB ePVS and MSNA **a** and **b** BG ePVS volume (mm3) with MSNA BF (bursts/min) and BI (bursts/100 heartbeats) **c** and **d** MB ePVS volume (mm3) with MSNA BF (bursts/min) and BI (bursts/100 heartbeats) **e** and **f** WM ePVS volume (mm3) with MSNA BF (bursts/min) and BI (bursts/100 heartbeats) **g** and **h** HP ePVS volume (mm3) with MSNA BF (bursts/min) and BI (bursts/100 heartbeats n=75 (25 hypertensive patients 50 controls)*p<0.05

Multiple linear regression analyses of the MSNA as a dependent variable were performed to determine if ePVS was associated with MSNA when adjusted for confounding factors such as age, sex, BP, and TIV. The results of the pooled analysis (n=75: n_hypertensive_=25,n_normotensive_=50) (Table S1) revealed that MSNA BI was associated with age, hypertension status, the number of ePVS clusters in WM and MB, TIV, and total WM volume. Moreover, MSNA BF was associated with age, sex, volume of WM ePVS, number of WM clusters as well as the volume of BG ePVS and MB ePVS.

The regression analysis for the hypertensive group (n=25, Table S2) revealed that MSNA BI was associated with age, BG ePVS volume, HCP ePVS volume, MB ePVS cluster count and TIV. Moreover, MSNA BF in the hypertensive group was associated with age, WM ePVS volume, WM ePVS cluster count, MB ePVS cluster count, TIV, and total WM volume.

The regression analysis of the normotensive group (n=50, Table S3) showed that MSNA BI and BF was associated with age, sex, WM ePVS volume, WM ePVS cluster, and BG ePVS volume. Also, MSNA BI in the normotensive group was positively correlated to the HP ePVS cluster counts.

Multiple linear regression was also performed on haemodynamic measurements as the dependent variable to determine the relationship between ePVS and haemodynamic measurements after adjusting for age, sex and MSNA. The pooled analysis of all participants (Table S4) revealed positive associations between WM ePVS volume with MBP and DBP. A negative association was observed between the WM ePVS cluster with, DBP, and PP. SBP was negatively associated with MB ePVS volume. In the hypertensives (Table S5), HP ePVS volume was positively associated with all haemodynamic measurements. SBP and PP showed a negative association with WM ePVS volume and a positive association with WM ePVS cluster. A similar pattern was observed in MB ePVS volume which showed a negative association with SBP and PP, and a positive association between MB ePVS cluster with SBP and PP. BG ePVS volume was negatively associated with DBP. In the normotensives (Table S6), SBP was negatively associated with the BG ePVS volume, TIV and positively associated with total WM volume. DBP was positively associated with WM ePVS volume but negatively associated with WM ePVS cluster counts.

## Discussion

Previous studies have suggested that ePVS is an emerging biomarker for a variety of disease states ^5,25,26^. How ePVS contribute to the pathophysiology has remained inconclusive. Here we show that one potential mechanism through which ePVS affects disease progression may be via the sympathetic nervous system. We have shown, for the first time, that MRI-visible ePVS are associated with MSNA. After adjusting for age, sex, MBP and TIV using multiple linear regression, we observed that MSNA in the hypertensive group was associated with ePVS in the WM, BG, HP, and MB, while the MSNA in the normotensive group was associated with ePVS in the WM, BG, and HP, but not in the MB. Our finding that enlarged PVS is associated with increased sympathetic nerve activity suggests that ePVS may contribute to the increased sympathetic vasoconstrictor drive in neurogenic hypertension.

### Potential mechanisms for the association between ePVS and sympathetic nerve activity

There are several ways ePVS may affect sympathetic outflow. Enlarged PVS are associated with changes in grey matter volume in cortical regions such as the orbitofrontal cortex, putamen and caudate ^27^, which are known brain regions that can modulate sympathetic activity ^22,28,29^. We also know that altered grey matter volume may affect resting levels of MSNA ^18,19^, suggesting it may be possible that ePVS disrupts sympathetic outflow via alteration of grey matter volumes. Interestingly, in our results, we observed that the hypertensive group showed a positive association between the midbrain ePVS and MSNA, which was not observed in the normotensive group. The midbrain contains key structures, such as the periaqueductal gray (PAG), known to modulate sympathetic nerve activity ^30^. It has been shown that electrical stimulations of PAG in humans may reduce MSNA ^31^. It could be hypothesised that disruption of the midbrain structures, such as PAG due to ePVS, could contribute to reduced output from this region, compromising the inhibitory effect of PAG on MSNA, which may contribute to elevated sympathetic drive in hypertension.

Moreover, another potential mechanism for the association between ePVS and MSNA in hypertension may be due to the disruption of astrocytes. Astrocytes, a key component that forms the perivascular space ^32,33^, can elevate sympathetic nerve activity ^34^. It has been shown that optogenetic stimulation of astrocytes in the rostral ventral lateral medulla (RVLM), the primary output nucleus for the generation of sympathetic vasoconstrictor drive, can elevate sympathetic nerve activity ^34^. Moreover, astrocytes in healthy states are sensitive to changes in cerebral perfusion pressure and can activate catecholaminergic sympathoexcitatory neurons to restore perfusion pressure to normative levels ^15^. In hypertension, however, it is known that astrocytes are perturbed ^35^, losing their function to serve as glial limitans that maintain the blood-brain barrier, leading to the penetration of leukocytes into the parenchyma^36^. Moreover, as part of the adaptive immune inflammatory response, astrocytes transition into their reactive state to form a physical barrier against leukocytes from entering the parenchyma ^37^. It is suggested that ePVS occur due to the loss of astrocytes’ ability to maintain fluid homeostasis via aquaporin 4 channel dysfunction, which can lead to impaired synaptic transmissions ^36,38^. Thus, it may be argued that disturbed astrocytes along perivascular spaces may underlie the abnormal sympathetic outflow in hypertension.

### Relationship between ePVS and blood pressure

Our results revealed no statistical differences between the normotensive and hypertensive groups in regard to total ePVS volume and cluster in the BG, HP, and MB and the ePVS volume in the WM. In fact, to our surprise, the total number of ePVS clusters in WM was significantly reduced in the hypertensive group compared to the normotensive group. This was possibly due to the reduction of the total WM and grey matter (GM) volume observed in the hypertensive group compared to the normotensive group. This hypothesis is supported by a previous study which reported that WM ePVS is associated with total intracranial volume^39^. To adjust for this confounding factor, we undertook further analyses for TIV, total WM volume, and total GM volume in all participants. After adjusting for these factors, we found that the hypertensive group showed a positive relationship between the number of ePVS clusters in WM with SBP and PP, which was not observed in the controls. This is consistent with the previous work which showed that cumulative BP exposure was positively associated with increases in WM ePVS score ^3^. Moreover, we found that WM ePVS volume was inversely correlated to SBP and PP, which may indicate that the hypertensives may have a higher number of ePVS clusters with smaller volumes. Indeed an intensive systolic blood pressure treatment has been shown to reduce ePVS volume ^40^. Therefore, it may be possible that the antihypertensive medication use in the hypertensive group may have influenced the WM ePVS volume. Moreover, there seems to be a mechanism that may limit the increases in the WM ePVS cluster in the normotensive group as we observed a negative association between the DBP and WM ePVS cluster after adjusting for confounding factors. This may be an important consideration for further work, noting that WM ePVS, specifically at the centrum semiovale, is associated with β-amyloid in Alzheimer’s disease patients ^41^.

### Limitations

One limitation of this study is its relatively small sample size. A larger sample may yield more statistically robust results, as structural MRI measures of brain regions exhibit high population variabilities ^42^. It is suggested that neuroimaging studies often underestimate the sample size required to generate statistically robust results and should consider using more than a thousand participants ^42^. However, given the invasive nature of recording sympathetic nerve activity via microneurography, and the time taken to obtain suitable data, undertaking such a large study would be impractical. Another potential confounding factor is individual differences in TIV, which may have influenced the amount of PVS detected. Therefore, we have adjusted all further analyses with the individual participant TIV, total WM volume, and total GM volume as mentioned previously.

Our investigation utilised two separate 3T MRI scanners from different manufacturers. Although the differences in voxel size were accounted for during the process of segmentation and pre-processing of the images, differences in resolution may have affected our results.

Moreover, using higher resolution scanners such as 7T scanners may enhance resolution and may yield more accurate detection of enlarged PVS in humans ^43^. Furthermore, MRI-visible ePVS are restricted to macroscopically visible enlarged spaces, which may not represent the entire perivascular network, such as pericapillary or perivenular spaces ^9,44^. Also, it has been suggested that PVS may be better visualised on the T2 weighted sequence than the T1 weighted sequence ^45^. However, we utilised an automated ePVS detection model optimised for T1-weighted images obtained from different scanners^23^, that enhances the accuracy of ePVS detection in the current dataset.

The MSNA burst analysis was limited to standard metrics such as BI and BF. Other complex measures of sympathetic nerve activity exist, including the continuous wavelet transform analysis which allows the assessment of the number of clusters of action potentials generated by individual axons from a multi-unit recording of MSNA ^46^, or single-unit recordings of MSNA ^47,48^. Nonetheless, with the relatively simple metrics and given a reasonable sample size, we were able to identify the significant association between ePVS and MSNA.

### Perspective

The long-standing limitation in applying microneurography relating to its time-consuming nature; the procedure ranges between 1.5 to 3 hours. Therefore, it is an impractical technique in clinical settings. Our results suggest the potential use of MRI-visible ePVS as an imaging biomarker for individuals with high sympathetic activity. Such an imaging marker may assist in identifying individuals who require microneurographic assessment of sympathetic nerve activity.

## Conclusions

Our results have shown an association between enlarged perivascular space and muscle sympathetic nerve activity in healthy adults and hypertensive individuals. While correlations between MSNA and ePVS were observed in the WM, BG and HP in both groups, the correlation between the MB ePVS and MSNA was only observed in the hypertensive group. This relationship provides insights into the pathophysiological mechanism of ePVS in hypertension.

## Data Availability

Data are available on reasonable request

## Acknowledgement

The authors acknowledge the support of research participants involved in the study. D.R wrote the article. D.R and W.P performed data analysis. D.R, R.F, L.H, and V.M obtained data. D.R, L.H and V.M designed the experiment. M.S, and A.H provided patients enrolled in the study.

## Sources of Funding

This work was funded by grants from the National Health and Medical Research Council of Australia to VGM and LAH (GTN1007557) and to VGM, AH and LAH (GTN1100042). DR was supported by an Australian Government Research Training Program (RTP) scholarship.

## Disclosures

M.P. Schlaich has received support from an National Health and Medical Research Council (NHMRC) Research Fellowship and research support from Medtronic, Abbott, and Servier Australia. He serves on scientific advisory boards for Abbott, Boehringer Ingelheim, Servier, Novartis, and Medtronic.

## Abbreviations

AHE: Adaptive histogram equalisation
BF: Burst frequency
BG: Basal ganglia
BI: Burst incidence
BOLD: Blood oxygen level dependent
CAT: Computational anatomy toolbox
DMH: Dorsomedial hypothalamus
ECG: Electrocardiogram
ePVS: Enlarged perivascular spaces
GM: Grey matter
HP: Hippocampus
MB: Midbrain
MRI: Magnetic resonance imaging
MSNA: Muscle sympathetic nerve activity
OSA: Obstrutive sleep apnoea
PAG: Periaqueductal gray
PVS: Perivascular spaces
RMS: Root mean squared
RVLM: Rostral ventrolateral medulla
SPM: Statistical parametric mapping
T: Tesla
T1: Longitudinal relaxation time
TE: Echo time
TIV: Toral intracranial volume
TR: Repetition time
WM: White matter

## Notes

### Competing Interest Statement

The authors have declared no competing interest.

### Clinical Trial

N/A

### Author Declarations

This cross-sectional study was approved by the Western Sydney University Human Research Ethics Committee (HREC approval H11462) and endorsed by Governance at the Baker Heart and Diabetes Institute.

